# Assessment of the Youth Empowerment Program on Mental Health Outcomes for Newcomer Youth in Aurora, CO – Lutheran Family Services Rocky Mountain

**DOI:** 10.1101/2025.10.18.25338030

**Authors:** Kiok Kim, Cole Young, Mary Jay, Jason Hwang, Jaewoo Seo, Hannah Lee, Christopher Park

## Abstract

**Background:** Mental health challenges among U.S. youth have become a leading cause of disability and poor life outcomes. Refugee and migrant youth face additional risks due to trauma experienced before, during, and after migration, compounded by discrimination and acculturation challenges. This project aims to assess the impact of the Youth Empowerment Program (YEP) as a mental health promotion plan for newcomer (US arrival within the past five years) youth in Aurora.

**Method:** This study utilized a pretest-posttest quasi-experimental design. 66 newcomer students participated in 20 weekly sessions focused on social-emotional skills and positive goal setting. Mental health outcomes were assessed using five validated self-report questionnaires measuring anxiety, depression, quality of life, resilience, and self-esteem at baseline and post-intervention. Data were analyzed using multiple linear regression by SAS 9.4.

**Results:** After the 20-week YEP intervention, 66 newcomer youth (Mean = 15) significantly improved mental health outcomes. Anxiety and depression decreased significantly (p <.0001), with females, Black students, and middle schoolers showing greater change. Quality of life improved (p =.0002), especially among Afghan students. Self-esteem increased, with Black students reporting the largest improvements. Resilience also improved significantly (p <.0001), with middle school students showing slightly better outcomes. The high effect sizes (R^2^ > 0.5) indicate a strong program impact.

**Conclusion:** The YEP effectively enhanced mental health outcomes for newcomer youth in Aurora, CO. Despite promising results, limitations such as no control group, convenience sampling, uneven subgroup sizes, and short-term follow-up suggest the need for future randomized studies with longer-term and qualitative evaluation.

## BACKGROUND

### Current mental health conditions among the young population in the US

Mental health challenges are common among children and youth and they have become the leading cause of disability and poor life outcomes in young people in the US. In recent years, significant increases have been detected in certain mental health disorders in youth, including depression, anxiety, and suicidal ideation (HHS, 2021). In the 2013-2019 data from CDC’s Mental Health Surveillance Among Children, among children and adolescents aged 12–17 years, 20.9% had ever experienced a major depressive episode. Among high school students in 2019, 36.7% reported persistently feeling sad or hopeless in the past year, and 18.8% had seriously considered attempting suicide. Approximately seven in 100,000 persons aged 10–19 years died by suicide in 2018 and 2019 (Bitsco et al., 2022). According to the Mental Health America report in 2018, Colorado ranked 48th out of 51 states in youth prevalence of mental illness, meaning Colorado youth have a higher prevalence of mental illness and lower rates of access to care. 11.93% of national youth (12-17) and 13.73% of Colorado youth reported mental issues and the number of youth experiencing major depressive episodes continues to rise annually at the national and state levels (Mental Health America, 2018). Along with mental health indicators, Colorado youth ranked 51st out of 51 states for the rate of youth with alcohol dependence and illegal drug use. National youth rates have decreased over time; however Colorado youth continued to show a high rank in substance use (Mental Health America, 2018). During the COVID-19 pandemic, many investigators reported elevated levels of depression or depressive symptoms, anxiety, stress or distress, externalizing behaviors, self-harm, and suicidal ideation and behaviors compared to the pre-pandemic period among the youth population (Zolopa et al., 2022). Colorado adolescents ages 11 to 18 have seen their rate of poor mental health double since 2017, from 8.8% to 18.5% in 2021 (Colorado Health Institute, 2023). Many factors shape the mental health of young people, from individual to societal level forces. It is especially important to protect the mental health of minority and marginalized young people. Due to factors beyond their control, these groups are at a higher risk for mental health difficulties and substance use (HHS, 2021).

### Current mental health conditions among refugee and migrant youth in the US

Worldwide, about 43.3 million children have been displaced as a consequence of conflict and violence as of the end of 2022, and increasing numbers of refugee and migrant youth are resettling in the US (UNICEF, 2023). In 2022, 25,465 refugees arrived in the US, and more than 40% were 18 and under (USA Facts, 2023). Refugees and migrants populations are exposed to extreme life conditions at different phases of displacement that affect their mental health and well-being: pre-migration (i.e., war, famine, torture, job and property loss, rape); during transit (i.e., family separation, physical and sexual assault, extortion, lack of access to services for basic needs); and post-migration (i.e., discrimination, acculturation shock, separation from family, detention, poor living conditions, barriers to accessing care) (WHO, 2023). Many researchers confirmed that refugee and migrant youth are at increased risk for depression, anxiety, PTSD, behavior problems, and social difficulties compared to children born in the US (Bunn & Betancourt, 2022) because their physical, emotional, and mental safety is not guaranteed amid racism, limited access to health care, and difficulties accessing legal, financial, and educational services (Mattar & Gellatly, 2022).

### Youth Empowerment Program (YEP)

Last year, in 2024, about 5000 refugees and 3000 migrants arrived in Colorado, and they were concentrated in the Denver-Metro area (Arapahoe, Denver, Broomfield, Jefferson, Douglas, and Adams counties). Among them, about 40% of the newcomer population was 18 or under (CDHS, 2025). Since they typically relocate to communities with high poverty, low resources, and a lack of academic and socio-emotional support in their schools, they have a higher chance of struggling with integration barriers such as language, acculturation, discrimination, and education gaps (Rodriguez, 2019). The Youth Empowerment Program (YEP) at the LFSRM is formulated to provide social-emotional learning and positive goal-setting skills to the newcomer youth population in the Denver Metro area. The YEP curriculum includes Cognitive Behavior Therapy (CBT) and Dialectical Behavior Therapy (DBT) and both approaches are based on Social Cognitive Theory (SCT). Its main focus is to improve the self-efficacy of refugee and migrant youth by regulating their feelings, thoughts, and actions and managing the diverse pressures in the school setting. Self-efficacy is the core construct of the SCT and it is believed to affect whether people will think positively or negatively, or choose to persevere when facing obstacles (Tip et al., 2020). YEP is a part of community efforts to promote mental health among the target population. According to Bandura (Bandura, 1998), comprehensive approaches that involve school-based health programs with familial and community efforts are more successful in promoting health than if schools try to do it alone. In the YEP curriculum, ‘Supporting Transition Resilience of Newcomer Groups (STRONG) for schools’ is incorporated as a CBT tool. STRONG for schools is an evidence-informed and school-based intervention for newcomer youth (K-12th) to support their transition to a new school and community (Hoover et al., 2019). The core components of STRONG for schools include resilience-building skills, understanding and normalizing distress, cognitive behavioral intervention skills, journey (before and after migration) narrative, and peer/parent/educator support (Hoover et al., 2019). According to Crooks et al, the major changes in the psychiatric outcomes by STRONG for schools are promoting resilience and reducing distress among refugee and immigrant students (Crooks et al., 2019; Crooks et al., 2020). DBT focuses on self-destructive behaviors to decrease life-threatening acts and increase mindfulness and self-management. It has demonstrated its benefit in stabilizing and controlling suicidal and parasuicidal behaviors (Panos et al., 2014). According to a meta-analysis (DeCou et al., 2019), DBT significantly reduces self-directed violence and use of psychiatric crisis services. However, DBT does not show a significant effect on reducing suicidal thoughts. These results suggest that DBT’s emphasis on managing behaviors over thoughts may influence its effectiveness and have implications for how it’s used in treating acute suicidality. The combination of CBT and DBT approaches warrants balancing changes in negative thoughts and destructive behaviors among newcomer students. DBT components in the YEP curriculum include mindfulness, emotion regulation, interpersonal effectiveness, and distress tolerance (Rathus & Miller, 2014). Therefore the purpose of this project is to assess the impact of the YEP as a mental health promotion plan for refugee and migrant youth in the Denver Metro area.

## METHODS

### Study Design

This study is designed as a one-group pretest-posttest quasi-experiment and administered in two schools - Aurora West College Preparatory School (6-12) and Gateway High School. Newcomer participants were recruited from August to September 2023 and were required to submit a parent/guardian consent form to their counselors. They were asked to complete five psychiatric self-report questionnaires to evaluate their mental health status in the first week (baseline; pretest) and the 20th week (endpoint; posttest) during the YEP in schools. All surveys were administered by the author, school counselors, and the school liaison. The score changes in the five scales were evaluated for intervention impacts. This project was approved or exempted through the Colorado School of Public Health; therefore, it was determined to be exempt from IRB review as a program evaluation.

### YEP School Groups

YEP facilitated a group session for newcomer youth in their schools for 20 weeks according to the curriculum. Each group had 15 to 20 individuals, and each participant filled out the student information form for their demographic data at the beginning of the group. Inclusion criteria for the YEP were refugee and migrant youth from other cultures, youth with a US arrival of less than five years (newcomer criteria), and middle and high school students in the Aurora public school district. A total of 82 participants were included in the program. However, 10 of them were excluded from the data analysis due to their suicidal ideation and behaviors on the PHQ-9A Depression scale. They were connected with the therapists after baseline data collection.

### Data Collection

As part of the assessment, each newcomer student was asked to answer five questionnaires at the first and 20th weeks of YEP. The five clinically validated self-report questionnaires are to measure (1) Anxiety, (2) Depression, (3) Quality of Life, (4) Resilience, and (5) Self-Esteem. Anonymized data was evaluated to detect any changes in five scales.

### Measures

1. Independent measures - Demographics: Each student was asked about their age, spoken language, US arrival time, home country, and how satisfied they were with their school.
2. Outcome measures - (1) Anxiety: Anxiety symptoms are measured using the Hospital Anxiety and Depression Scale (HADS) (White et al., 1999). For this study, seven questions in the Anxiety domain are included. Questions are answered on a Likert scale ranging from 0–3, with the answers changing depending on the question. Scores ranged from 0–21, with higher scores indicating a greater degree of anxiety (Reeson et al., 2020). (2) Depression: Depression symptoms are measured using the Patient Health Questionnaire - Adolescent version (PHQ-A) (Richardson et al., 2010). Each question is scored on a Likert scale that ranges from 0 (not at all) to 3 (nearly every day). Scores range from 0–27, with higher scores indicating a greater degree of depression (Reeson et al., 2020). (3) Quality of Life: Quality of Life is measured by using the KIDSCREEN-10 (Ravnes-Sieberer et al., 2010). Each question is scored on a Likert scale that ranges from 0 (not at all/never) to 4 (extremely/always). Scores range from 0–44, with higher scores indicating a better quality of life (Reeson et al., 2020). (4) Resilience: Resilience is measured using the Child & Youth Resilience Measure – adolescent version (CYRM-12) (Govender et al., 2017). Answers range from 1 (Not at all) to 5 (A lot), and scores range from 12–60, with higher scores indicating more characteristics associated with resilience (Reeson et al., 2020). (5) Self-Esteem: Self-Esteem is measured using the Rosenberg Self-Esteem Scale (RSES) (Bagley & Mallick, 2001). Each question is scored on a Likert scale (0–3) that ranges from Strongly Agree to Strongly Disagree, depending on the question. Scoring is achieved through summation of results and can range from 0–40, with higher scores indicating better self-esteem (Reeson et al., 2020).

### Statistical Analysis

To compare mean scores at baseline and endpoint, multiple linear regression was carried out by using SAS 9.4. For all statistical tests, p<0.05 is considered a statistically significant change in survey and question scores. Results show the mean and standard deviation.

## RESULTS

Of the 82 students initially enrolled, 66 of them were included in the data analysis. As in Table 1, the mean age of participants was 15.0 years (SD=1.8). In terms of gender distribution, 37 students (56%) identified as male and 29 students (44%) identified as female. The racial and ethnic composition of the sample included 36 students (55%) who identified as Afghan Caucasian, 16 students (24%) who identified as Black, and 14 students (21%) who identified as Hispanic. Students represented a diverse range of nationalities, including Somalia, South Sudan, Congo, Rwanda, Uganda, Guinea, Tanzania, Burundi, Mexico, Colombia, Guatemala, Honduras, Venezuela, Peru, and Afghanistan.

**Table 1.** Demographics of newcomer students.

|  |  |  |
| --- | --- | --- |
| Number of students |  | 66 |
| Mean age ± SD | | 15 ± 1.8 |
| Gender | Male | 37 (56%) |
|  | Female | 29 (44%) |
| Race/Ethnicity | Afghan Caucasian | 36 (55%) |
|  | Black | 16 (24%) |
|  | Hispanic | 14 (21%) |
| Grade | Middle | 17 (26%) |
|  | High | 49 (74%) |

After 20 weekly sessions of social emotional learning, participants reported statistically significant improvements across all five measured domains (Tables 2 and 3). Anxiety scores, as measured by HADS-A, decreased significantly (p<0.0001), with females demonstrating greater reductions than males. Depression scores, assessed via PHQ9-A, also decreased significantly (p<0.0001), with Black students showing greater improvements compared to Hispanic students. Quality of life, measured by KIDSCREEN-10, improved significantly (p=0.0002), with Afghan students reporting slightly higher gains than Hispanic students. Resilience scores, assessed by CYRM-12, improved significantly (p<0.0001), with middle school students demonstrating marginally greater gains compared to high school students.

**Table 2.** Mean change in self-report outcome measures after 20-week of YEP (n = 66)

| Questionnaire | Domain | Range | Mean (Baseline) | Mean (Post) | $\Delta$ | p | $R^2$ |
| --- | --- | --- | --- | --- | --- | --- | --- |
| HADS | Anxiety | 0–21 | 6.6 | 5 | –1.6 | <0.0001* | 0.82 |
| PHQ9-A | Depression | 0–27 | 7.9 | 5.9 | –2 | <0.0001* | 0.88 |
| KIDSCREEN-10 | Quality of Life | 0–44 | 24.4 | 26.1 | 1.7 | 0.0002* | 0.55 |
| CYRM-12 | Resilience | 12–60 | 46 | 48.8 | 2.8 | <0.0001* | 0.56 |
| RSES | Self-Esteem | 0–40 | 22.7 | 23.9 | 1.2 | <0.0001* | 0.79 |

**Table 3.** Demographic groups’ intervention effects across measures.

| **Domain** | **Gender**<br>(Female and Male) | **Race/Ethnicity**<br>(Hispanic, Black, Afghan) | **Grade Group**<br>(50 High: 16 Middle) |
| --- | --- | --- | --- |
| Anxiety | Very significant ($p < .0001$)<br>Female > Male | Not significant | Not significant |
| Depression | Not significant | Significant ($p = .0013$)<br>Black > Hispanic | Significant ($p = .0083$)<br>High > Middle |
| Quality of Life | Not significant | Marginal ($p = .0589$)<br>Afghan > Hispanic | Not significant |
| Resilience | Not significant | Not significant | Marginal ($p = .0564$)<br>Middle > High |
| Self-Esteem | Not significant | Significant ($p = .0444$)<br>Black > Afghan | Not significant |

Similarly, self-esteem, as evaluated by the Rosenberg Self-Esteem Scale, showed substantial improvement (p<0.0001), particularly among Black students. Especially, among racial and ethnic groups, Black students showed greater improvement in depression and self-esteem domains. This finding suggests that the YEP sessions may have a greater positive impact on Black students compared to their Afghan and Hispanic peers. The effect sizes for all outcomes were strong (R^2^ > 0.5), suggesting a substantial impact of the YEP intervention on mental health outcomes.

## DISCUSSION

The findings from this study demonstrate that the Youth Empowerment Program (YEP) had a significant and positive impact on the mental health of newcomer youth in the Aurora Public School District. Statistically significant improvements across anxiety, depression, quality of life, resilience, and self-esteem domains indicate that the intervention successfully addressed critical aspects of students’ emotional and psychological well-being. YEP conveyed several strengths. First, the integration of CBT and DBT approaches within a culturally sensitive, school-based framework allowed the program to meet the complex needs of refugee and migrant youth. The inclusion of diverse activities such as peer tutoring, field trips, job training, and cultural celebrations enhanced participant engagement and the overall sense of community, which are essential components of mental health promotion among displaced populations. Furthermore, the use of validated outcome measures and rigorous statistical analysis adds to the robustness of the findings. The program demonstrated a strong intervention impact, as evidenced by high effect sizes (R^2^ > 0.5). This indicates that a substantial proportion of the variance in outcomes can be attributed to the intervention. Finally, the inclusion of Afghan, Black, and Hispanic youth from diverse countries enhances the project’s efforts toward health equity and culturally responsive programming. However, several limitations should be addressed. The absence of a control group limits the ability to attribute observed improvements solely to the YEP intervention and could be a threat to internal validity. In addition, the use of convenience sampling and the relatively small sample size may limit the generalizability of results to broader newcomer youth populations in other school settings. The unequal distribution between subgroups may have introduced biases in subgroup analyses. Also, the study’s short-term follow-up captures only immediate post-intervention effects and doesn’t guarantee long-term sustainability of the intervention impacts on mental health improvements. Therefore, future studies should prioritize the implementation of randomized controlled trials to find real causal inferences regarding intervention effects. Longitudinal designs incorporating at least six- and twelve-month follow-up assessments are necessary to evaluate the durability of observed improvements. Incorporating qualitative methods, such as key informant interviews with school counselors/school liaisons and focus groups, would also provide valuable insights into how students experienced the program and how to improve the program in the future. Lastly, scaling up the YEP model across additional schools and regions could contribute to a broader movement toward culturally responsive mental health promotion for refugee and migrant youth in broader communities.

### PUBLIC HEALTH SIGNIFICANCE OF THE PROJECT

The Youth Empowerment Program (YEP) provided critical mental health support for newcomer youth in Aurora CO, leading to significant improvements in anxiety, depression, quality of life, resilience, and self-esteem. This finding contributes valuable evidence to the field of public health by demonstrating the effectiveness of a culturally responsive and school-based mental health intervention program tailored to refugee and migrant youth, a marginalized population in traditional mental health systems. By integrating CBT and DBT strategies with community-centered supports such as peer tutoring and cultural events, YEP addresses mental health through a health equity perspective. The program’s success highlights the importance of embedding mental health services within accessible school settings for youth navigating the multiple challenges of acculturation and education gaps. The program impacts underscore the need for scalable and inclusive public health strategies that are trauma-informed and community-engaged approaches to enhance public health’s commitment to improving mental well-being and reducing disparities among vulnerable youth populations.

## Data Availability

All data produced in the present work are contained in the manuscript

## DECLARATIONS

### Ethical Approval

This project was reviewed as part of the author’s Master of Public Health capstone at the Colorado School of Public Health and was determined to be exempt from institutional review board (IRB) as a program assessment.

### Competing Interest Statement

The author declares no competing interests. This project was completed as part of the author’s Master of Public Health capstone at the Colorado School of Public Health. The author received no payments or financial support from any third party or organization.

## Acknowledgments

The author thanks Lutheran Family Services Rocky Mountain and the Aurora Public School District for their collaboration and support of this project. Especially, she thanks Mary Jay, MSW, for her mentorship and supervision during the program assessment, Cole Young for his assistance in group facilitation and data collection, and peer tutors for their efforts and constructive feedback in teaching and mentoring newcomer students. Lastly, she thanks all participants in the YEP sessions at Aurora Public Schools.

## Notes

### Competing Interest Statement

The authors have declared no competing interest.

### Funding Statement

This study did not receive any funding

### Author Declarations

Ethics Committee/IRB of the Colorado School of Public Health waived the ethical approval for this work.

